# Beyond green cover: Greenspace morphology and configuration predict heat-related illness in Arizona

**DOI:** 10.64898/2026.07.08.26357485

**Authors:** Huaqing Wang, Shujuan Li, Simin Gholami, Joseph Hoover, Mackenzie Waller, Kacey Ernst

## Abstract

Residential greenness has been associated with reduced heat-related illness, yet the specific role of greenspace morphology at the neighborhood scale remains insufficiently understood. This study quantified the relationship between heat-related illness and multiple dimensions of greenspace morphology using an eight-year (2016–2023) unbalanced panel dataset comprising 19,021 block-group–year observations across 2,427 census block groups in Arizona, USA. One meter high-resolution National Agricultural Imagery Program aerial imagery was classified to calculate greenspace percentage, number of greenspaces, average size, shape complexity, connectedness, and distantness, at the block group level. We applied conditional spatial autoregressive models with a negative binomial distribution to estimate associations between each morphology metric and yearly heat-related illness counts, adjusting for sociodemographic and geographic covariates. We found higher greenspace percentage, aggregation, shape complexity, connectedness, and density were consistently associated with lower heat-related illness risk. A one–standard deviation increases in shape complexity corresponded to a 12.4% decrease in expected heat-related illness counts (IRR = 0.876, 95% CI: 0.834–0.921). Similarly, increases in greenspace percentage (14.6% decrease; IRR = 0.855, 95% CI: 0.827–0.885), number of greenspace patches (3.7% decrease; IRR = 0.963, 95% CI: 0.937–0.990), average size (4.5% decrease; IRR = 0.955, 95% CI: 0.923–0.989), and connectedness (5.5% decrease; IRR = 0.945, 95% CI: 0.918–0.972) were all protective. In contrast, larger inter-greenspace distances were associated with increased heat-related illness risk (6.1% increase; IRR = 1.061, 95% CI: 1.033–1.091). Our findings highlight the critical importance of multiple dimensions of greenspace morphology in mitigating heat-related health risks. These results suggest that heat-reduction planning with greening initiatives should consider not only the amount of greenspace but also its spatial configuration to maximize cooling and result in health benefits.

**Plain Language Summary:** Extreme heat is a growing health risk, especially in hot regions such as Arizona. While greener neighborhoods are known to reduce heat-related illnesses, less is known about whether the *arrangement* of green spaces also matters. In this study, we examined how different features of neighborhood green space—such as how much green space there is, how large it is, how connected it is, and how far apart green areas are—relate to heat-related illness over time. Using eight years of data from more than 2,400 neighborhoods across Arizona, we found that neighborhoods with more green space, greener areas that are closer together, better connected, and more evenly distributed experienced fewer heat-related illnesses. In contrast, neighborhoods where green spaces were sparse and far apart had higher risks of heat-related illness. These findings show that reducing heat-related health risks is not just about adding more green space, but also about designing greener neighborhoods in smarter ways. Urban heat-reduction strategies can be more effective when they consider both the amount of greenery and how green spaces are arranged within communities.

## Introduction

Extreme heat is a rapidly intensifying public health threat worldwide, with heat-related morbidity and mortality increasing sharply in recent decades (eClinicalMedicine, 2024; WHO, 2024). Urban areas bear a disproportionate burden of such heat related health risk due to the combined effects of dense built environments, limited vegetation, and the intensification of urban heat islands (Heaviside et al., 2017; Hsu et al., 2021a; Zheng et al., 2026). In heat-vulnerable regions, such as the southwestern United States, reducing the rising burden of heat-related illness requires identifying environmental determinants that mitigate extreme-heat exposure (Howard et al., 2024; Narayanan & Keellings, 2025).

A growing body of evidence indicates that urban greenness plays an important role in reducing ambient temperatures, and, in turn, lowering population exposure to extreme heat and subsequent heat-related illness (Lin & Li, 2025; Nazish et al., 2024a; Wong et al., 2021). A recent global modeling study demonstrated that vegetation substantially reduce heat-related mortality and that greening is a key climate-adaptation strategy (Wu et al., 2025). Importantly, this work also highlighted that the protective effects of greenness depend on both its presence *and* spatial configuration, calling for research that explicitly examines the characteristics and distribution of greenspace. A recent systematic review on greenspace morphology and health also reported a lack of literature in greenspace morphology and heat related health risks (Wang, Gholami, et al., 2024).

Despite substantial research, the cooling effects of vegetation field still lack consensus on whether larger clustered vegetation or smaller dispersed greenery provides greater protection against heat-related health risks. This uncertainty is driven in part by methodological differences, temperature metrics, and spatial scales. In terms of cooling capacity, one study reported that the magnitude of cooling and transport (how far it extends into surrounding areas) depends on the size, spread, and geometry of greenspaces. Certain large, solitary parks show limited cooling beyond the immediate surface boundary layer (Gunawardena et al., 2017). Others studies suggested that targeted, larger vegetation clusters outperform dispersed greenery (Fan et al., 2015; Probst et al., 2022; Wang et al., 2023). For related health outcomes, it has been reported that small, accessible parks embedded within residential areas may yield the greatest reduction in heat-related mortality (He et al., 2026). Recent epidemiological evidence suggests that landscape-level characteristics, such as higher coupling, connectivity, and less complex boundary shapes, reduce heat-mortality risks (Yao et al., 2026a). Together, these conflicting findings underscore that greenspace morphology is likely consequential, yet its specific role in shaping population vulnerability to heat remains insufficiently resolved.

Key limitations of the existing literature include the use of coarse spatial resolution (>10 m) to characterize greenspace and reliance on city-level metrics that obscure neighborhood-scale exposure variability. In greenspace and health outcome association studies, very few analyses employ high-resolution (e.g.,1 m) imagery to measure urban greenspace morphology and even fewer evaluate multiple, spatially dependent dimensions of landscape configuration. Consequently, it remains unclear which specific configuration characteristics of greenspace - its shape, number, connectedness, or inter-greenspace distance - most strongly influence heat-related illness in the urban environments.

To address these gaps, we used an eight-year spatial panel dataset of 2,427 neighborhoods in Arizona, combining 1 meter NAIP aerial imagery with greenspace morphology metrics to quantify relationships between six dimensions of landscape configuration and heat-related illness. This study advances our understanding of how urban neighborhood-level greenspace morphology shapes heat-related health risks by leveraging high-resolution environmental data with a spatial autoregressive model in Arizona, where heat exposure ranges from moderate (high altitude north) to extreme (south and central). A clear understanding of greenspace morphology may directly inform heat-mitigation planning at the neighborhood scale where interventions are often most feasible.

## Methods

### Study area and heat related illness

Analyses were conducted at the census block group level in the state of Arizona, United States. Arizona is among the most heat-vulnerable states in the U.S., characterized predominantly by arid and semi-arid climates with hot summers (>40°C) and low annual precipitation (<30 cm) in the central and southern portions of the state where the majority of the population resides. The state also experiences a disproportionately high burden of heat-related health outcomes compared to other U.S. states. Between 2013 and 2024 heat deaths increased from 151 deaths in 2013 to nearly 977 (Arizona Department of Health Services, 2024).

To focus on contexts where the built environment characteristics are most strongly linked to heat exposure and intervention opportunities, we restricted the study area to block groups located within census-defined urbanized areas with a density equal or above 425 units per square mile (U.S. Census Bureau, 2023). This restriction reflects the evidence that the urban heat island effects due to impervious surfaces, anthropogenic heat and lower vegetation increase heat exposure and heat-related illnesses. Urban greenspace is also more directly shaped by planning and municipal planning making them better targets for evaluating modifiable landscape characteristics impacts on health.

We obtained statewide emergency department (ED) visits and hospital discharge records from the Arizona Department of Health Services for the years 2016–2023, which included each patient’s primary and secondary diagnoses coded according to the International Classification of Diseases, Tenth Revision (ICD-10). Heat-related illness cases were identified using ICD-10 codes T67, which encompasses clinical heat-related conditions such as heat stroke, heat exhaustion, heat cramps, and other specified heat effects, and X30, which denotes exposure to excessive natural heat. These two codes are widely used in prior epidemiological research on heat-related morbidity and mortality (Dring et al., 2022; Harduar Morano & Watkins, 2017; Petitti et al., 2016). Following case identification, patient residential addresses were geocoded via Google Geocoder API in R, and annual counts of heat-related illness events were spatially aggregated to the corresponding census block groups via ArcGIS Pro to support subsequent geographic and statistical analyses.

### Greenspace image classification

We mapped greenspace using high-resolution aerial imagery from the US Geological Survey National Agriculture Imagery Program (NAIP), which provided one-meter resolution multispectral images for 2015, 2017, 2019, and 2023; the imagery was collected during the leaf-on period and with cloud cover below 10%. Vegetation was identified using a vegetation-index thresholding approach, in which each year’s imagery was processed to separate vegetated from non-vegetated pixels based on a calibrated vegetation index cutoff (Hashim et al., 2019; Huang et al., 2021; Wang et al., 2026; Wang & Tassinary, 2024). Classification performance met standard remote-sensing expectations, with overall accuracy above 90% and Kappa values exceeding 0.80, indicating satisfactory accuracy (Morales-Barquero et al., 2019). The image classification and accuracy assessment were conducted using Google Earth Engine.

### Quantification of residential greenspace morphology

We delineated the surrounding local environment by generating a 0.5-mile radial buffer around each block group. This approach accounts for residents living near block group edges who may interact more frequently with greenspaces located in adjacent block groups than within their own. A distance of 0.5 miles approximates a 10-minute walking radius and has been widely adopted in prior environmental exposure and accessibility studies (Browning & Lee, 2017; Wang et al., 2024; Wang & Tassinary, 2019, 2024). This buffer size is particularly appropriate for evaluating heat-related health impacts, as recent evidence indicates that 0.5-mile buffers capture stronger associations than either smaller or larger spatial scales (Song et al., 2024). Given the considerable computational demands of high-resolution greenspace morphology calculations, we restricted the analysis to a 0.5-mile buffer and did not assess additional spatial radii. All buffer generation and spatial processing were performed using Python and ArcGIS Pro.

Within each census block group’s buffered landscape, we quantified six morphological characteristics of local greenspace. These metrics capture different dimensions of landscape composition and pattern, including the overall proportion of vegetated land, the number and size distribution of greenspace patches, geometric complexity, and the degree to which patches are spatially connected or isolated. Specifically, we derived measures representing overall greenspace coverage (PLAND), number of greenspaces (NP), average size (AREA_MN), shape complexity (SHAPE_AM), landscape cohesion (COHESION), and mean nearest-neighbor distance (ENN_MN). A visual overview of these metrics is provided in Figure 1, and formal definitions and computational procedures, based on established landscape ecology practices as documented in the Fragstats methodological literature (McGarigal, 2014; McGarigal et al., 2012), are summarized in Table 2.

**Figure 1.**
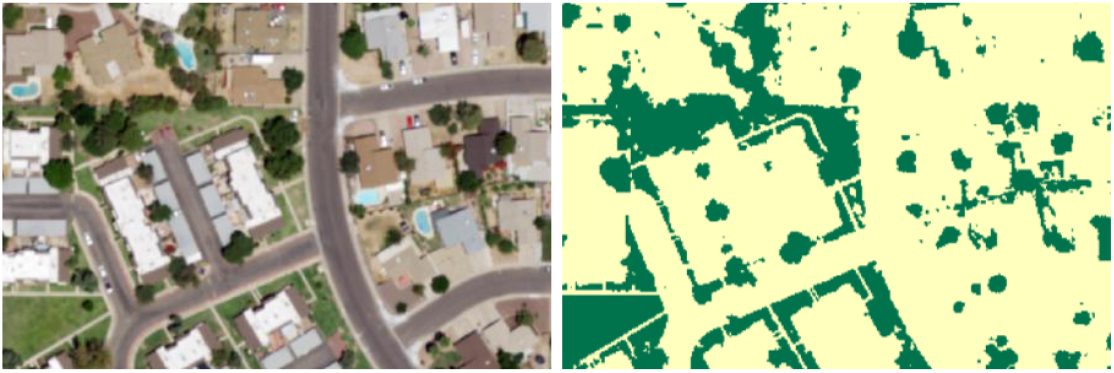
Example of image classification results (Left: NAIP satellite imagery. Right: classified greenspace map)

**Figure 2.**
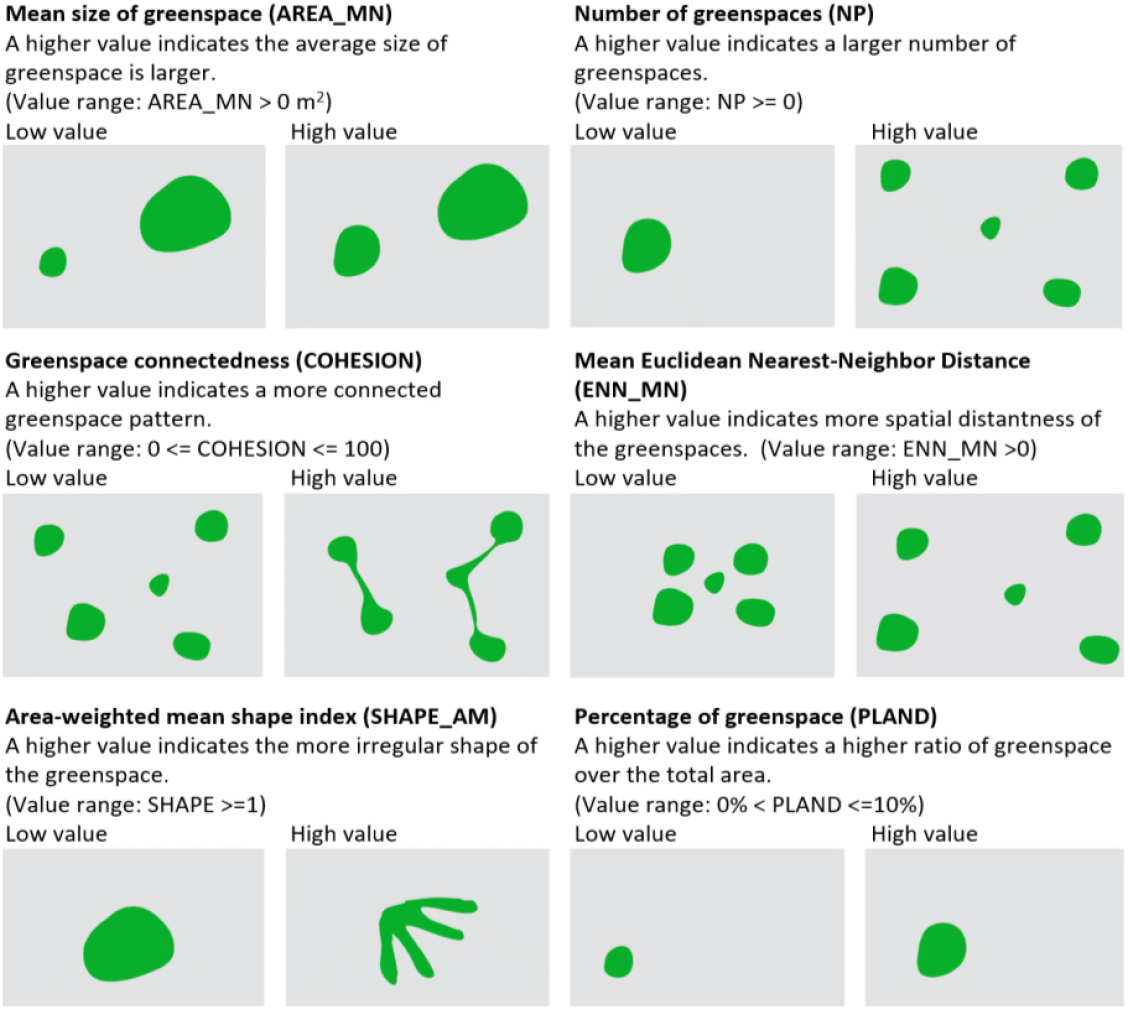
Illustration of Greenspace Morphology Metrics (Some images in this table are adopted from a previous paper *(H. Wang* & *Tassinary, 2024)* with additional images generated by the authors of the current study for the purpose of enhancing data presentation. Copyright 2024 Huaqing Wang. Used with permission.)

**Figure 3.**
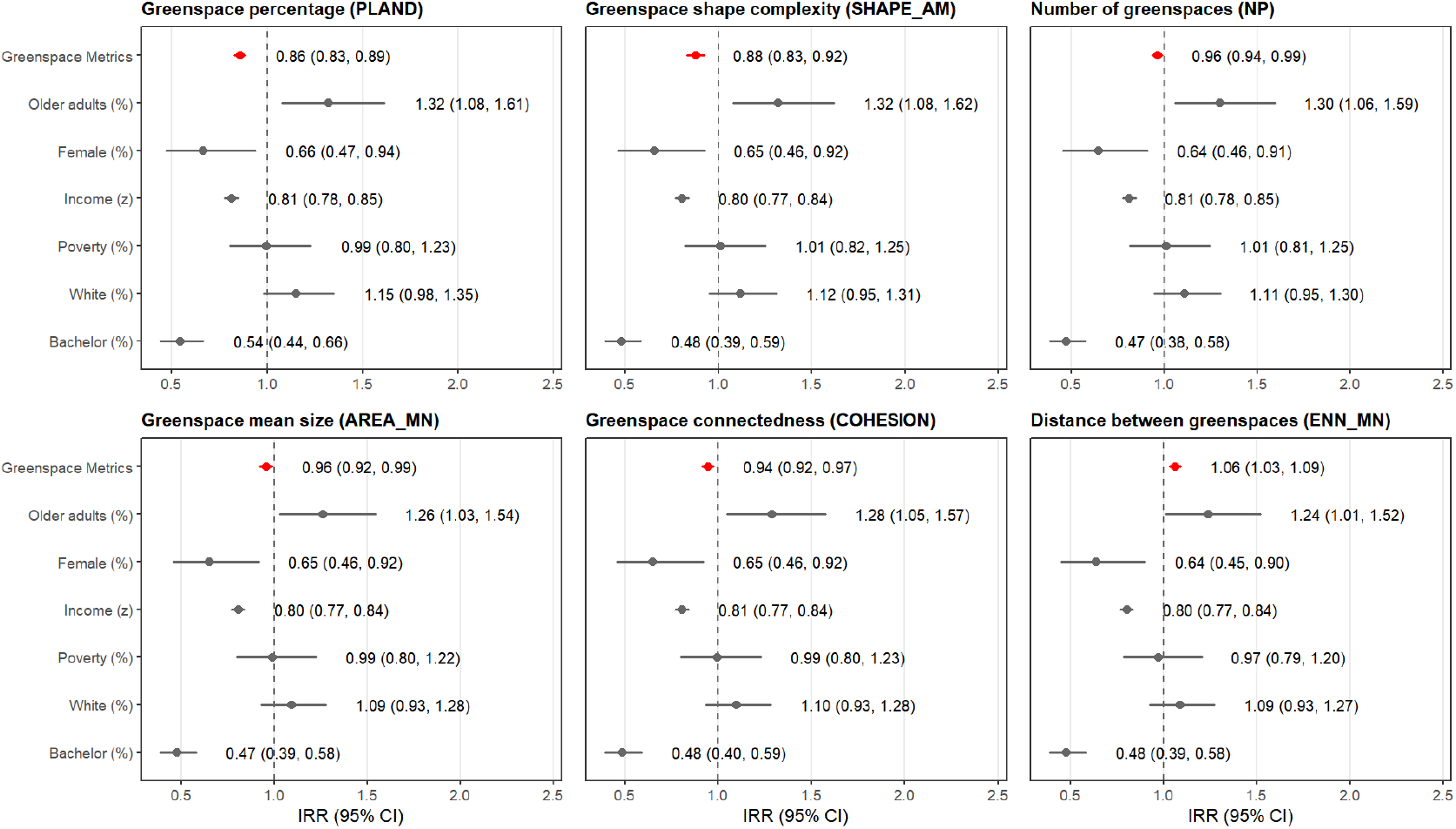
Forest plots of incidence rate ratios (IRRs) associating greenspace morphology metrics to heat-related illness across Arizona block groups, 2016–2023. Red points denote the focal greenspace metric; gray points denote covariates; bars indicate 95% CIs. Effects correspond to a one–standard deviation increase in each metric; models adjust for sociodemographic covariates, year fixed effects, spatial adjacency, and a log(population) offset.

The selection of greenspace morphology indicators was informed by three overarching criteria. First, we prioritized metrics that have demonstrated empirical validity in prior environmental health and landscape epidemiology research, particularly those identified in a recent systematic review as having consistent and meaningful associations with human health outcomes (Shen & Lung, 2016, 2017; Wang, Gholami, et al., 2024; Wang & Tassinary, 2019). Second, we incorporated spatial characteristics that have been identified as key determinants of the thermal performance of urban vegetation, features consistently highlighted in studies examining how greenspace configuration modulates cooling intensity and distribution (Guo et al., 2019; Kowe et al., 2021a; Y. Li et al., 2023; C. Wang et al., 2019; Zhou et al., 2011). These properties are directly relevant to understanding population vulnerability to heat. Finally, we selected metrics that provide interpretable and actionable insights for urban planners and policymakers, ensuring that resulting findings can meaningfully inform landscape design and climate-adaptation strategies.

In years lacking aerial imagery coverage, we estimated greenspace morphology metric values by applying a spline-based temporal interpolation using data from adjacent imagery years. For instance, morphology values for 2016 were derived by interpolating between the corresponding 2015 and 2017 measurements for the same census block group. Because greenspace morphology typically changes gradually over time in established urban environments, especially at the neighborhood scale, such interpolation introduces limited bias and is widely considered acceptable for high-resolution longitudinal environmental exposure assessment (Bai et al., 2023; Liu et al., 2024). All morphology computations were carried out using the *landscapemetrics* package (Hesselbarth et al., 2019) in R.

### Covariates

We incorporated a set of sociodemographic and economic indicators measured at the census block group level to account for neighborhood characteristics that may confound the relationship between greenspace patterns and heat-related illness. These variables were obtained from the U.S. Census Bureau’s American Community Survey (ACS) and merged by block group and year. Heat vulnerable populations include the elderly, individuals who cannot afford to cool their homes, individuals with disabilities, and peoples of color historically marginalized (Gronlund, 2014; Hsu et al., 2021b). The selected indicators represent these dimensions by including proportion of older adults (aged ≥65), the percentage of female residents, the percentage of white residents, median household income, the share of adults holding a bachelor’s degree or higher, the percentage of individuals living below the federal poverty line, and total population size.

### Statistical analysis

We evaluated the relationship between greenspace morphology and heat-related illness using a conditional spatial autoregressive model specified with a negative binomial distribution. This modeling framework is well suited for over dispersed count outcomes, consistent with the distributional properties observed in the heat-related illness data. Prior to model selection, we assessed spatial dependence in the residuals using Moran’s I (Supplementary Table S3), which revealed significant positive spatial autocorrelation (p < 0.001), indicating that a spatially explicit regression approach was necessary. To construct a time-invariant spatial weights matrix, we excluded block groups with boundary changes between 2016 and 2023 (N=4632, 19.6%), and such changes typically reflect substantial population increases. These boundary changes alter adjacency relations and introduce potential confounding from population increase. Restricting the sample to block groups with stable boundaries ensured consistent comparative neighborhood boundaries with a more demographically stable analytic population. After applying this criterion and removing observations with missing data, the analytic dataset consisted of 19,021 block-group–year observations from 2,427 block groups across 2016–2023.

Because several greenspace morphology metrics capture related dimensions of landscape configuration and therefore exhibit moderate intercorrelation (Table 2), we modeled each metric separately to avoid instability related to multicollinearity and to estimate its association with heat-related illness without adjustment for the other morphology measures. All models incorporated the neighborhood-level covariates described above. To facilitate comparison across predictors measured on different scales and to improve numerical stability and model convergence, all greenspace morphology metrics were standardized prior to regression analysis. VIFs were tested for each model, and were all found below 4, indicating minimum level of collinearity. All analyses were conducted using *spaMM* package in R (Rousset & Ferdy, 2014).

## Results

Across block-group–year observations, the mean count of heat-related illness was 0.793 (SD 1.839; range 0–90), indicating a sparse outcome with occasional high counts. Greenspace morphology showed substantial heterogeneity: the percentage of greenspace averaged 17.44% (SD 6.37), shape complexity averaged 4.60 (SD 4.77), and landscapes were generally fine-grained and well-connected (mean number of greenspaces 5518.50; mean connectedness 94.83; mean nearest-neighbor distance 3.82). Sociodemographic composition was diverse, with mean percentage of older-adult of 18.4%, female percentage of 51%, and average median household income of $68,524 (Table 1).

**Table 1.**
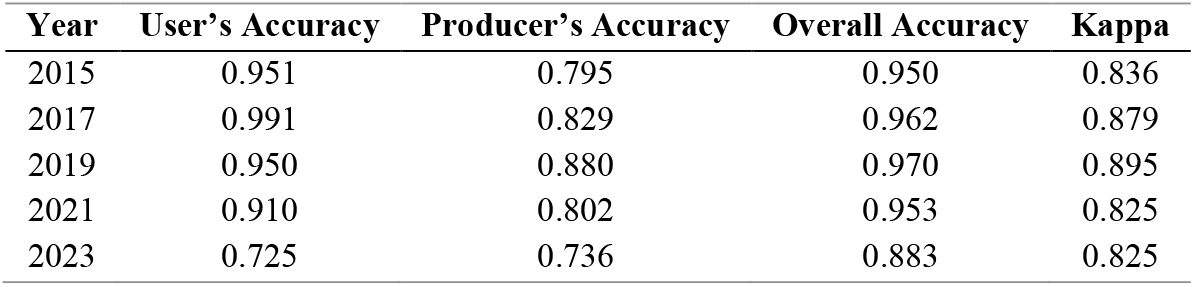
The threshold values of the Normalized Difference Vegetation Index (NDVI) used in the image classification process, along with the corresponding accuracy of the greenspace classification.

| Year | User's Accuracy | Producer's Accuracy | Overall Accuracy | Kappa |
| --- | --- | --- | --- | --- |
| 2015 | 0.951 | 0.795 | 0.950 | 0.836 |
| 2017 | 0.991 | 0.829 | 0.962 | 0.879 |
| 2019 | 0.950 | 0.880 | 0.970 | 0.895 |
| 2021 | 0.910 | 0.802 | 0.953 | 0.825 |
| 2023 | 0.725 | 0.736 | 0.883 | 0.825 |

The pairwise correlation analysis suggests that greener block groups tended to have more aggregated and connected greenspace (Table 2). Greenspace percentage correlated positively with connectedness (r = 0.713) and average greenspace size (r = 0.597), and negatively with nearest-neighbor distance (r = −0.497). Income closely aligned with education (r = 0.638) and inversely with poverty (r = −0.603). Correlation coefficients with heat-related illness were small but directionally consistent. The coefficient is lower where greenness coverage was higher/connected (e.g., greenspace % r = −0.104; connectedness r = −0.076) and higher where green patches were farther apart (nearest-neighbor distance r = 0.049); heat-related illness also decreased with the increases of income (r = −0.123) and education (r = −0.149) and increased with poverty increase (r = 0.149).

A total of 19,021 block-group–year observations from 2,427 census block groups (2016–2023) were included in the analysis. All models were conditional spatial autoregressive models with negative binomial distribution, including year fixed effects and sociodemographic adjustment.

**Table 2.** Detailed formula for calculating landscape pattern metrics*.

| Landscape indices (acronym) | Measured Morphological Characteristic | Formula (unit and range) | Description |
| --- | --- | --- | --- |
| Percentage of greenspaces (PLAND) | Greenspace percentage | $PLAND = \frac{\sum_{j=1}^n a_{ij}}{A} * 100$<br>(Value range: 0% < PLAND <= 100%) | $a_{ij}$ = the area of each patch.<br>$A$ = the total landscape area.<br>•A higher value indicates a higher ratio of greenspace over the total area. |
| Mean size of greenspaces | Mean Size of Greenspace | $AREA = a_{ij} \left( \frac{1}{10000} \right), MN = \frac{\sum_{j=1}^n x_{ij}}{n_i}$<br>(hectares: AREA_MN > 0, without limit) | $a_{ij}$ = area (hectares) of patch $ij$.<br>$MN$ = the sum, across all patches of the corresponding patch type, of the corresponding patch metric values, divided by the number of patches of the same type.<br>•A higher value indicates a larger average size of greenspaces, and vice versa. |
| Number of Patches (NP) | Number of Greenspaces | $NP = n_i$<br>(Number of greenspaces; NP > 0) | $n_i$ = number of patches in the landscape of patch type (class) $i$.<br>•A higher value denotes a denser distribution of greenspace, while a lower value suggests the opposite. |
| Greenspace Cohesion Index (COHESION) | Greenspace connectedness | $COHESION = \left( 1 - \frac{\sum_{j=1}^n p_{ij}}{\sum_{j=1}^n p_{ij} \sqrt{a_{ij}}} \right) \left( 1 - \frac{1}{\sqrt{A}} \right)^{-1} (100)$<br>(None; 0 < COHESION < 100) | $p_{ij}$ = perimeter of patch $ij$ in terms of the number of cell surfaces.<br>$a_{ij}$ = area of patch $ij$ in terms of the number of cells.<br>$A$ = total number of cells in the landscape.<br>•A lower value indicates that greenspaces become progressively subdivided and less physically connected. |
| Area weighted mean shape index (SHAPE_AM) | Greenspace spatial shape complexity | $SHAPE = \frac{p_{ij}}{\min(p_{ij})}, AM = \sum_{j=1}^n \left[ x_{ij} \left( \frac{a_{ij}}{\sum_{j=1}^n a_{ij}} \right) \right]$<br>(None; SHAPE > 1, without limit.) | $p_{ij}$ = perimeter of patch $ij$ in terms of the number of cell surfaces.<br>$\min(p_{ij})$ = minimum perimeter of patch $ij$ in terms of the number of cell surfaces.<br>$AM$ (area-weighted mean) equals the sum, across all patches of the corresponding patch type, of the corresponding patch metric value multiplied by the proportional abundance of the patch [i.e., patch area (m<sup>2</sup>) divided by the sum of patch areas]. |
|  |  |  | •A higher value signifies a more complex shape of greenspace, and vice versa. |
| Mean of Euclidean nearest neighbor distance (ENN_MN) | Spatial distantness of greenspaces | $ENN = h_{ij}, MN = \frac{\sum_{j=1}^n x_{ij}}{n_i}$ (Meters, ENN > 0, without limit) | $h_{ij}$ = distance (m) from patch $ij$ to nearest neighboring patch of the same type (class), based on patch edge-to-edge distance, computed from cell center to cell center.<br>MN = the sum, across all patches of the corresponding patch type, of the corresponding patch metric values, divided by the number of patches of the same type.<br>•A higher value signifies a greater distance between greenspaces. |

\* Information in this table is referenced from the FRAGSTATS HELP manual (McGarigal, 2014).

**Table 3.** Descriptive statistics of outcomes, greenspace morphology, and covariates.

| Metrics | Mean | Std.Dev | Min | Max |
| --- | --- | --- | --- | --- |
| Heat related illness count | 0.793 | 1.839 | 0.000 | 90.000 |
| ***Greenspace Morphology Metrics*** |  |  |  |  |
| Percentage of greenspace (%) | 17.438 | 6.365 | 1.710 | 68.966 |
| Greenspace shape complexity | 4.597 | 4.774 | 1.565 | 260.561 |
| Number of greenspaces | 5518.497 | 6389.847 | 127.942 | 144035.425 |
| Average greenspace size | 37.916 | 19.244 | 4.106 | 474.166 |
| Greenspace connectedness | 94.828 | 2.552 | 77.668 | 100.237 |
| Nearest-neighbor distance between greenspaces | 3.824 | 0.580 | 2.435 | 7.846 |
| ***Social Demographic Factors*** |  |  |  |  |
| Older adults (%) | 0.184 | 0.186 | 0.000 | 1.000 |
| Female (%) | 0.508 | 0.064 | 0.026 | 0.832 |
| Median household income | 68524.312 | 34452.657 | 4234.000 | 250001.000 |
| Poverty (%) | 0.149 | 0.135 | 0.000 | 0.849 |
| White (%) | 0.746 | 0.178 | 0.000 | 1.000 |
| Bachelor degree or higher (%) | 0.309 | 0.194 | 0.000 | 0.957 |
| Population size | 1559.991 | 664.602 | 88.000 | 5835.000 |

**Table 4.** Correlation coefficient matrix between heat related illness, greenspace morphology metrics, and social-demographic variables.

|  | 1 | 2 | 3 | 4 | 5 | 6 | 7 | 8 | 9 | 10 | 11 | 12 | 13 |
| --- | --- | --- | --- | --- | --- | --- | --- | --- | --- | --- | --- | --- | --- |
| 1 Percentage of greenspace (%) | 1 |  |  |  |  |  |  |  |  |  |  |  |  |
| 2 Greenspace shape complexity | 0.500 *** | 1 |  |  |  |  |  |  |  |  |  |  |  |
| 3 Number of greenspaces | 0.149 *** | 0.144 *** | 1 |  |  |  |  |  |  |  |  |  |  |
| 4 Average greenspace size | 0.597 *** | 0.357 *** | -0.230 *** | 1 |  |  |  |  |  |  |  |  |  |
| 5 Greenspace connectedness | 0.713 *** | 0.404 *** | -0.019 ** | 0.607 *** | 1 |  |  |  |  |  |  |  |  |
| 6 Nearest-neighbor distance | -0.497 *** | -0.093 *** | -0.011 | -0.023 ** | -0.252 *** | 1 |  |  |  |  |  |  |  |
| 7 Older adults (%) | 0.145 *** | 0.134 *** | 0.206 *** | 0.029 *** | 0.171 *** | 0.003 | 1 |  |  |  |  |  |  |
| 8 Female (%) | 0.047 *** | 0.041 *** | 0.033 *** | 0.012 | 0.049 *** | -0.015 * | 0.271 *** | 1 |  |  |  |  |  |
| 9 Median household income | 0.284 *** | 0.062 *** | 0.282 *** | 0.073 *** | 0.176 *** | -0.124 *** | -0.036 *** | -0.049 *** | 1 |  |  |  |  |
| 10 Poverty (%) | -0.238 *** | -0.058 *** | -0.196 *** | -0.095 *** | -0.202 *** | 0.124 *** | -0.247 *** | -0.085 *** | -0.603 *** | 1 |  |  |  |
| 11 White (%) | 0.167 *** | 0.046 *** | 0.112 *** | 0.177 *** | 0.138 *** | -0.029 *** | 0.442 *** | 0.112 *** | 0.179 *** | -0.318 *** | 1 |  |  |
| 12 Bachelor degree or higher (%) | 0.376 *** | 0.101 *** | 0.202 *** | 0.212 *** | 0.261 *** | -0.165 *** | 0.138 *** | 0.012 | 0.638 *** | -0.483 *** | 0.322 *** | 1 |  |
| 13 Heat related illness count | -0.104 *** | -0.041 *** | 0.003 | -0.072 *** | -0.076 *** | 0.049 *** | -0.067 *** | -0.066 *** | -0.123 *** | 0.149 *** | -0.123 *** | -0.149 *** | 1 |

\*\*\* indicates statistically significant at 0.001 level, \*\* indicates statistically significant at 0.01 level, and \* indicates statistically significant at 0.05 level

Across all six greenspace morphology metrics, higher greenness coverage and more favorable spatial configuration were consistently associated with lower heat-related illness risk, whereas greater separation among green patches was associated with higher risk. Specifically, a one–standard deviation (SD) increase in greenspace percentage (PLAND) was associated with a 14.5% reduction in expected heat-related illness incidence (IRR = 0.855; 95% CI: 0.827–0.885). Protective associations of similar magnitude were observed for higher greenspace shape complexity (IRR = 0.8761; 95% CI: 0.833–0.921), greater patch density (IRR = 0.963; 95% CI: 0.937–0.990), larger average greenspace patch size (IRR = 0.955; 95% CI: 0.923–0.989), and greater greenspace connectedness (IRR = 0.945; 95% CI: 0.918–0.972). In contrast, greater nearest-neighbor distance between greenspaces was positively associated with heat-related illness (IRR = 1.061; 95% CI: 1.033–1.091). Effect sizes were highly consistent across models (Figure 1; Supplementary Materials Tables S1–S6).

Sociodemographic gradients were stable in magnitude and direction across all six models. Higher proportions of older adults were positively associated with heat related illness risk (IRRs 1.24–1.32 across models). Higher median household income (IRRs ≈0.80) and greater proportion of residents with bachelor level educational attainment (IRRs ≈0.47–0.54) were consistently protective. Percentage of female was inversely associated with heat-related illness. We did not observe any association with the heat related illness risk for the percentage of residents under poverty line and the percentage of White residents.

Year fixed effects (Supplementary Materials Tables S4–S9) indicated pronounced temporal increases in heat-related illness risk relative to 2016. The largest increases were observed in 2022 and 2023, when incidence was approximately 1.5-fold and 2-fold higher, respectively, independent of greenspace morphology and sociodemographic factors.

## Discussion

Using high resolution greenspace maps and an eight-year spatial panel covering more than 2,400 neighborhoods in one of the most heat-vulnerable U.S. states, this study provides compelling evidence that both the amount and the morphology of greenspace play critical roles in shaping population vulnerability to heat-related illness. Across all six morphological dimensions, we identified highly consistent protective associations: neighborhoods with higher percentage of green coverage, contained greater number of green patches, and exhibited more aggregated, connected, and complex vegetated configuration experienced lower heat-related illness counts, whereas neighborhoods with greater distances between greenspace patches were associated with elevated risk. These findings underscore that greenspace is not merely an aesthetic or ecological amenity but a foundational component to build heat resilience in urban environments.

### Greenspace percentage and shape

Our finding that neighborhoods with a higher percentage of greenspace experience lower heat-related illness risk is consistent with evidence from broader spatial scales. A recent systematic review similarly reported that regions with more abundant greenspace show reduced rates of heat-related morbidity compared with sparsely vegetated areas (Nazish et al., 2024b).

Our finding that more complex greenspace shapes are associated with lower heat-related illness risk added additional perspectives, compared with several prior studies reporting that simpler park geometries were protective against heat-related mortality at larger spatial scales (He et al., 2026; Yao et al., 2026b). These complementary findings highlight the possibility of scale dependence of greenspace–health relationships. These earlier studies were conducted at the city or subdistrict level, where analytic units span 5 to 20 square kilometers and contain populations of 50,000 to 150,000 residents (references). These studies also relied on relatively coarse land-cover data ranging from 10 to 30 meters resolution. At this resolution the “shape” of greenspaces captured largely reflects the outlines of large greenspaces/parks and does not accurately capture the fine-scale geometry of neighborhood vegetation. Under such conditions, important greenery such as narrow vegetated strips, street-tree corridors, irregular edges, or small green land parcels cannot be resolved, and the resulting shape metrics describe only the morphology of large greenspaces/parks.

In contrast, our analysis used one meter resolution imagery to calculate greenspace configuration at the census block-group scale, where the mean spatial unit area is 0.96 square kilometers and the mean population is 1,560 residents. At this finer resolution, greenspace shape captures micro-scale vegetation features that penetrate into residential areas, increase edge interfaces, create localized shading, and support small pockets of evaporative cooling. These characteristics are more likely to exhibit complex forms, and their cooling benefits are directly experienced by residents, particularly during extreme heat. The seemingly divergent findings across studies therefore reflect differences in both spatial scale and data resolution rather than true contradictions. Coarse-scale analyses capture the macro-level geometry of large greenspaces/parks, where simpler boundaries may facilitate airflow and cooling, while high-resolution neighborhood analyses capture small, vegetated elements whose irregularity and embeddedness enhance thermal mitigation. Together, these results emphasize that greenspace– health relationships are highly scale-dependent and that the spatial resolution of greenspace data must align with the ecological and human-exposure processes under examination.

### Greenspace numbers and size

Results from this study suggest that neighborhoods containing a greater number of greenspaces experienced lower heat-related illness risk, even when individual greenspaces were small. This pattern supports widespread tree-planting and micro-greening initiatives by demonstrating that vegetation patches as small as one square meter can contribute measurable cooling benefits when sufficiently numerous. At the same time, we observed that larger average greenspace size was protective. The mean greenspace patch size in our dataset was 38 square meters, reflecting a landscape dominated by very small, vegetated elements such as individual street trees, narrow verge plantings, and small parcels embedded within parking lots and along roadways. Our results therefore suggest that expanding the size of these small patches can substantially amplify cooling benefits and further reduce heat-related illness risk. Together, these results indicate that both micro-greening and the expansion of small, vegetated elements may provide meaningful thermal benefits.

These findings are consistent with a growing body of literature on greenspace cooling performance. A prior study showed that park area was a positive predictor of cooling intensity and cooling distance in a cross-city analysis, indicating that larger parks provide more powerful and spatially extensive thermal mitigation (Tian et al., 2025). Similarly, across 41 cities in the Yangtze River Delta, a study reported that larger blue–green space areas were strongly associated with lower land-surface temperatures, demonstrating that extensive vegetated patches produce more substantial regional cooling than smaller or fragmented ones (X. Li et al., 2025). Together, these results reinforce the importance of both increasing the number of greenspaces and strategically expanding their size to maximize their cooling potential and protect populations from heat-related illness.

### Greenspace connectedness and inter-greenspace distance

Through this study, we also observed that higher greenspace connectedness and shorter distances between greenspaces were each associated with reduced heat-related illness risk at the neighborhood level. These results suggest that tree-planting programs and greening investments that strengthen vegetative continuity or reduce gaps between patches may enhance local cooling and lower heat-related illness risk. This interpretation is supported by prior work demonstrating that the spatial cohesion of greenspace is a key determinant of cooling performance. A prior study reported that more aggregated and better connected vegetation produced stronger reductions in land-surface temperature across urban functional zones (Q. Wang et al., 2023). Beyond thermal regulation, greenspace connectivity has also been linked with lower risks of non-communicable disease mortality and morbidity, reduced preterm birth, healthier childhood body mass index, improved mental health, higher cancer survival, and lower myopia prevalence among school-age children (H. Wang, Gholami, et al., 2024, 2024; H. Wang, Huang, et al., 2024; H. Wang & Tassinary, 2019). Together, these findings highlight the importance of considering not only the quantity of greenspace but also the continuity and spatial configuration of vegetated areas when designing interventions aimed at reducing heat-related health risks.

### Potential mechanisms of cooling by greenspace characteristics

We believe that the association between greenspace morphology and heat-related illness is primarily driven by the additional cooling benefits and the accessibility provided by specific spatial characteristics of vegetation, beyond the influence of greenspace abundance alone. Previous studies have shown that clustered vegetation reduces surface temperatures more effectively than dispersed vegetation, suggesting that spatial aggregation enhances thermal performance (Chen et al., 2024). However, spatial aggregation of greenspace may result in a smaller number of greenspace patches and lower accessibility to greenspaces. Other morphological features, including patch size, vegetation density, shape complexity, and spatial connectedness, have also been found to influence the magnitude and spatial reach of cooling (Upreti & Kumar, 2025; Xie et al., 2013; Zhou et al., 2011). Among these characteristics, connectedness consistently demonstrates the strongest inverse relationship with land-surface temperature, highlighting the importance of continuous or closely spaced vegetated patches in sustaining localized cooling (Kowe et al., 2021b). These mechanisms offer a plausible explanation for the consistent associations we observed across multiple morphological dimensions. Taken together, these mechanisms emphasize that the spatial arrangement of vegetation plays an essential role in determining its cooling capacity and offers valuable guidance for planners and policymakers seeking to maximize the health benefits of urban greening.

### Limitations

Several limitations should be considered when interpreting these findings. First, our greenspace classification did not distinguish among vegetation types such as trees, shrubs and grassland, largely due to the challenges of separating these categories using high-resolution aerial imagery in arid environments. Given that trees generally provide stronger shading and evaporative cooling than low-stature vegetation, future work with enhanced spectral or LiDAR data would clarify vegetation-type-specific mechanisms. Second, although our study benefits from very fine spatial resolution, the analysis was conducted entirely within the unique climatic and development context of Arizona, which encompasses substantial heterogeneity—from cooler, high-elevation regions in the north to hot desert environments in the central and southern parts. Neighborhoods in many study areas are characterized by arid-adapted vegetation and distinct heat exposure profiles, which may limit the generalizability of our findings to cities with markedly different climates, vegetation structures, or built-environment configurations. Third, our panel design does not account for shorter variations in behavioral adaptations at the individual level.

High-frequency mobility data may allow more direct assessment of exposure pathways in future studies. Finally, although spatial models helped address autocorrelation and isolate associations with greenspace morphology, unmeasured confounders such as irrigation practices, shade from buildings, or micro-scale cooling infrastructure may still influence results. Continued research integrating detailed environmental measurements, individual-level exposure data and multi-city comparisons will be essential for validating and extending these findings.

## Conclusion

This study provides robust evidence that the amount and the spatial configuration of greenspace play an important role in shaping vulnerability to heat-related illness in one of the most heat-exposed regions of the United States. By integrating high-resolution aerial imagery with an eight-year neighborhood-level spatial panel, we demonstrate that several dimensions of greenspace morphology, including patch abundance, average patch size, shape complexity, connectedness and interpatch distance, contribute meaningfully to heat-health outcomes.

Neighborhoods that were greener, more structurally connected and composed of larger or more intricately shaped vegetation patches consistently showed lower risks of heat-related illness, while greater separation among vegetated patches was associated with higher risk.

These findings highlight the importance of considering not only how much greenspace is available in urban environments but also its spatial arrangement. Efforts that increase the number of vegetated patches, expand small greenspaces, improve connectivity through green corridors, and reduce gaps between vegetated areas may enhance cooling at the neighborhood scale and help protect residents during extreme heat. As climate change intensifies the frequency and severity of dangerous heat events, optimizing greenspace morphology represents an actionable strategy for strengthening heat-health resilience in cities. Future work should assess whether these patterns hold in regions with different climates, vegetation types, and urban forms, and should evaluate the effectiveness of specific greening interventions using finer environmental measurements and individual-level exposure data. Continued research at varied spatial scales with multi-resolution greenspace maps, and diverse methodological approaches, will be essential for identifying the most impactful greening strategies. Our findings suggest that urban greening strategies that incorporate spatial configuration, rather than focusing on quantity alone, may offer the greatest potential for protecting public health under intensifying climate change.

## Data Availability

The heat-related illness data are not publicly available due to restrictions imposed by the Arizona Department of Health Services. The greenspace-related datasets generated during the current study are available from the corresponding author upon reasonable request.

## Open Research

The data used in this study were obtained from a combination of publicly available and restricted-access sources. Greenspace morphology metrics were derived from National Agricultural Imagery Program (NAIP) aerial imagery, which is publicly available through the U.S. Geological Survey (https://www.usgs.gov/centers/eros/science/usgs-eros-archive-aerial-photography-national-agriculture-imagery-program-naip). Socio-demographic covariates were obtained from the American Community Survey and are publicly accessible (https://www.census.gov/programs-surveys/acs.html). Heat-related illness data were obtained from the Arizona Department of Health Services (https://www.azdhs.gov/), and are subject to data use agreements that restrict public redistribution of individual-level health records; therefore, these data cannot be publicly shared. All statistical analyses were conducted using R, and the R code used for data processing is provided in the Appendix to support transparency and reproducibility.

