## Supplementary Materials for "Beyond green cover: Greenspace morphology and configuration predict heat-related illness in Arizona"

Table 1. Morean's I test results

| Model | Moran's I | P-value |
|---|---|---|
| PLAND | 4.1093 | 1.985e-05 |
| NP | 4.3468 | 6.906e-06 |
| AREA_MN | 4.3573 | 6.583e-06 |
| SHAPE | 4.2154 | 1.247e-05 |
| COHESION | 4.3103 | 8.15e-06 |
| ENN_MN | 3.7731 | 8.063e-05 |

Table 2. Regression coefficients of greenspace shape complexity (SHAPE_AM) in predicting heat related illness

| Variable | Estimate | SE | t | IRR | 95% CI: Lower | 95% CI: Upper |
|---|---|---|---|---|---|---|
| Greenspace Shape Complexity (SHAPE_AM) | -0.132 | 0.025 | -5.192 | 0.876 | 0.833 | 0.921 |
| Percentage of Older Adults | 0.278 | 0.104 | 2.682 | 1.320 | 1.078 | 1.618 |
| Percentage of Female Residents | -0.424 | 0.177 | -2.403 | 0.654 | 0.463 | 0.925 |
| Median Household Income (z-score) | -0.217 | 0.022 | -9.853 | 0.805 | 0.770 | 0.840 |
| Percentage of Residents Under Poverty Line | 0.011 | 0.108 | 0.103 | 1.011 | 0.818 | 1.251 |
| Percentage of White Residents | 0.111 | 0.081 | 1.371 | 1.118 | 0.953 | 1.310 |
| Percentage of Residents with Bachelor's Degree or Higher | -0.734 | 0.103 | -7.162 | 0.480 | 0.393 | 0.587 |
| Year: 2017 | 0.096 | 0.036 | 2.688 | 1.101 | 1.026 | 1.181 |
| Year: 2018 | 0.074 | 0.036 | 2.071 | 1.077 | 1.004 | 1.155 |
| Year: 2019 | 0.058 | 0.036 | 1.614 | 1.060 | 0.988 | 1.138 |
| Year: 2020 | -0.043 | 0.038 | -1.116 | 0.958 | 0.889 | 1.033 |
| Year: 2021 | 0.108 | 0.039 | 2.780 | 1.114 | 1.032 | 1.201 |
| Year: 2022 | 0.383 | 0.039 | 9.836 | 1.467 | 1.359 | 1.583 |
| Year: 2023 | 0.680 | 0.040 | 16.892 | 1.975 | 1.825 | 2.137 |

Notes: IRR = incidence rate ratio. SE = conditional standard error. N = 19,021 block-group–year observations across 2,427 census block groups (2016–2023).

Table 3. Regression coefficients of greenspace percentage (PLAND) in predicting heat-related illness

| **Variable** | **Estimate** | **SE** | **t** | **IRR** | **95% CI: Lower** | **95% CI: Upper** |
|---|---|---|---|---|---|---|
| Greenspace Percentage (PLAND) | -0.156 | 0.017 | -8.97 | 0.855 | 0.827 | 0.885 |
| Percentage of Older Adults | 0.276 | 0.103 | 2.67 | 1.318 | 1.076 | 1.614 |
| Percentage of Female Residents | -0.411 | 0.176 | -2.33 | 0.663 | 0.469 | 0.937 |
| Median Household Income (z-score) | -0.210 | 0.022 | -9.50 | 0.811 | 0.777 | 0.847 |
| Percentage of Residents Under Poverty Line | -0.007 | 0.108 | -0.06 | 0.993 | 0.803 | 1.228 |
| Percentage of White Residents | 0.140 | 0.081 | 1.73 | 1.151 | 0.982 | 1.349 |
| Percentage of Residents with Bachelor's Degree or Higher | -0.614 | 0.104 | -5.92 | 0.541 | 0.442 | 0.663 |
| Year: 2017 | 0.106 | 0.036 | 2.96 | 1.111 | 1.036 | 1.192 |
| Year: 2018 | 0.070 | 0.036 | 1.96 | 1.073 | 1.000 | 1.150 |
| Year: 2019 | 0.055 | 0.036 | 1.52 | 1.057 | 0.984 | 1.134 |
| Year: 2020 | -0.049 | 0.038 | -1.28 | 0.952 | 0.883 | 1.026 |
| Year: 2021 | 0.028 | 0.040 | 0.71 | 1.029 | 0.951 | 1.113 |
| Year: 2022 | 0.378 | 0.039 | 9.73 | 1.460 | 1.353 | 1.575 |
| Year: 2023 | 0.726 | 0.041 | 17.83 | 2.068 | 1.909 | 2.239 |

Notes: IRR = incidence rate ratio. SE = conditional standard error. N = 19,021 block-group–year observations across 2,427 census block groups (2016–2023)

Table 4. Regression coefficients of greenspace patch density (NP) in predicting heat-related illness

| **Variable** | **Estimate** | **SE** | **t** | **IRR** | **95% CI: Lower** | **95% CI: Upper** |
|---|---|---|---|---|---|---|
| Greenspace Patch Density (NP) | -0.037 | 0.014 | -2.67 | 0.963 | 0.937 | 0.990 |
| Percentage of Older Adults | 0.262 | 0.104 | 2.51 | 1.300 | 1.059 | 1.595 |
| Percentage of Female Residents | -0.439 | 0.177 | -2.49 | 0.645 | 0.456 | 0.911 |
| Median Household Income (z-score) | -0.211 | 0.022 | -9.46 | 0.810 | 0.775 | 0.846 |
| Percentage of Residents Under Poverty Line | 0.007 | 0.109 | 0.06 | 1.007 | 0.814 | 1.246 |
| Percentage of White Residents | 0.104 | 0.081 | 1.28 | 1.110 | 0.946 | 1.301 |
| Percentage of Residents with Bachelor's Degree or Higher | -0.754 | 0.103 | -7.35 | 0.471 | 0.385 | 0.575 |
| Year: 2017 | 0.088 | 0.036 | 2.45 | 1.092 | 1.018 | 1.171 |
| Year: 2018 | 0.075 | 0.036 | 2.10 | 1.078 | 1.005 | 1.157 |
| Year: 2019 | 0.065 | 0.036 | 1.80 | 1.067 | 0.994 | 1.146 |
| Year: 2020 | -0.043 | 0.039 | -1.11 | 0.958 | 0.888 | 1.033 |
| Year: 2021 | 0.113 | 0.039 | 2.91 | 1.119 | 1.037 | 1.207 |
| Year: 2022 | 0.382 | 0.039 | 9.73 | 1.465 | 1.357 | 1.582 |
| Year: 2023 | 0.675 | 0.042 | 16.23 | 1.965 | 1.811 | 2.132 |

Notes: IRR = incidence rate ratio. SE = conditional standard error. N = 19,021 block-group–year observations across 2,427 census block groups (2016–2023).

Table 5. Regression coefficients of average greenspace patch size (AREA_MN) in predicting heat-related illness

| **Variable** | **Estimate** | **SE** | **t** | **IRR** | **95% CI: Lower** | **95% CI: Upper** |
|---|---|---|---|---|---|---|
| Greenspace Mean Area (AREA_MN) | -0.046 | 0.018 | -2.59 | 0.955 | 0.923 | 0.989 |
| Percentage of Older Adults | 0.231 | 0.104 | 2.23 | 1.260 | 1.028 | 1.544 |
| Percentage of Female Residents | -0.434 | 0.177 | -2.45 | 0.648 | 0.458 | 0.917 |
| Median Household Income (z-score) | -0.217 | 0.022 | -9.84 | 0.805 | 0.770 | 0.840 |
| Percentage of Residents Under Poverty Line | -0.010 | 0.109 | -0.10 | 0.990 | 0.800 | 1.225 |
| Percentage of White Residents | 0.086 | 0.081 | 1.05 | 1.089 | 0.929 | 1.277 |
| Percentage of Residents with Bachelor's Degree or Higher | -0.745 | 0.103 | -7.23 | 0.475 | 0.388 | 0.581 |
| Year: 2017 | 0.048 | 0.037 | 1.28 | 1.049 | 0.975 | 1.128 |
| Year: 2018 | 0.058 | 0.036 | 1.61 | 1.060 | 0.988 | 1.137 |
| Year: 2019 | 0.051 | 0.036 | 1.40 | 1.052 | 0.980 | 1.130 |
| Year: 2020 | -0.076 | 0.039 | -1.93 | 0.927 | 0.858 | 1.001 |
| Year: 2021 | 0.118 | 0.039 | 3.05 | 1.125 | 1.043 | 1.213 |
| Year: 2022 | 0.329 | 0.041 | 7.95 | 1.390 | 1.282 | 1.508 |
| Year: 2023 | 0.579 | 0.046 | 12.47 | 1.784 | 1.628 | 1.953 |

Notes: IRR = incidence rate ratio. SE = conditional standard error. N = 19,021 block-group–year observations across 2,427 census block groups (2016–2023).

Table 6. Regression coefficients of greenspace connectedness (COHESION) in predicting heat-related illness

| Variable | Estimate | SE | t | IRR | 95% CI: Lower | 95% CI: Upper |
|---|---|---|---|---|---|---|
| Greenspace Connectedness (COHESION) | -0.057 | 0.015 | -3.86 | 0.945 | 0.918 | 0.972 |
| Percentage of Older Adults | 0.250 | 0.104 | 2.41 | 1.285 | 1.048 | 1.575 |
| Percentage of Female Residents | -0.430 | 0.177 | -2.43 | 0.650 | 0.460 | 0.920 |
| Median Household Income (z-score) | -0.215 | 0.022 | -9.74 | 0.806 | 0.772 | 0.842 |
| Percentage of Residents Under Poverty Line | -0.007 | 0.109 | -0.06 | 0.993 | 0.803 | 1.229 |
| Percentage of White Residents | 0.091 | 0.081 | 1.12 | 1.095 | 0.934 | 1.284 |
| Percentage of Residents with Bachelor's Degree or Higher | -0.726 | 0.103 | -7.04 | 0.484 | 0.395 | 0.592 |
| Year: 2017 | 0.075 | 0.036 | 2.12 | 1.078 | 1.006 | 1.156 |
| Year: 2018 | 0.065 | 0.036 | 1.82 | 1.067 | 0.995 | 1.145 |
| Year: 2019 | 0.057 | 0.036 | 1.57 | 1.058 | 0.986 | 1.136 |
| Year: 2020 | -0.062 | 0.038 | -1.61 | 0.940 | 0.872 | 1.014 |
| Year: 2021 | 0.100 | 0.039 | 2.55 | 1.105 | 1.023 | 1.192 |
| Year: 2022 | 0.352 | 0.039 | 9.01 | 1.422 | 1.317 | 1.535 |
| Year: 2023 | 0.629 | 0.040 | 15.80 | 1.876 | 1.735 | 2.029 |

Notes: IRR = incidence rate ratio. SE = conditional standard error. N = 19,021 block-group–year observations across 2,427 census block groups (2016–2023).

Table 7. Regression coefficients of nearest-neighbor greenspace distance (ENN_MN) in predicting heat-related illness

| Variable | Estimate | SE | t | IRR | 95% CI: Lower | 95% CI: Upper |
|---|---|---|---|---|---|---|
| Greenspace Distantness (ENN_MN) | 0.060 | 0.014 | 4.31 | 1.061 | 1.033 | 1.091 |
| Percentage of Older Adults | 0.216 | 0.104 | 2.09 | 1.241 | 1.013 | 1.521 |
| Percentage of Female Residents | -0.449 | 0.177 | -2.54 | 0.638 | 0.451 | 0.902 |
| Median Household Income (z-score) | -0.222 | 0.022 | -10.05 | 0.801 | 0.767 | 0.837 |
| Percentage of Residents Under Poverty Line | -0.027 | 0.109 | -0.25 | 0.973 | 0.787 | 1.204 |
| Percentage of White Residents | 0.083 | 0.081 | 1.03 | 1.087 | 0.927 | 1.274 |
| Percentage of Residents with Bachelor's Degree or Higher | -0.740 | 0.103 | -7.21 | 0.477 | 0.390 | 0.583 |
| Year: 2017 | 0.129 | 0.038 | 3.42 | 1.137 | 1.057 | 1.224 |
| Year: 2018 | 0.075 | 0.036 | 2.10 | 1.078 | 1.005 | 1.156 |
| Year: 2019 | 0.076 | 0.036 | 2.10 | 1.079 | 1.005 | 1.158 |
| Year: 2020 | -0.042 | 0.038 | -1.09 | 0.959 | 0.889 | 1.034 |
| Year: 2021 | 0.041 | 0.043 | 0.95 | 1.042 | 0.958 | 1.133 |
| Year: 2022 | 0.384 | 0.039 | 9.83 | 1.469 | 1.360 | 1.586 |
| Year: 2023 | 0.715 | 0.043 | 16.58 | 2.045 | 1.879 | 2.225 |

Notes: IRR = incidence rate ratio. SE = conditional standard error. N = 19,021 block-group–year observations across 2,427 census block groups (2016–2023).

Table 8. R code for regression analysis

```
library(sp)
library(spdep)
library(spaMM)
library(sf)

bg <- st_read("D:/USU/NIH_ClimateHealth_Lab -
General/Heat_Illness_Study/1_StatisticalAnalysis/5_SpatialShapfile/tl_2020_04_bg/tl_2020_04
_bg.shp", quiet = TRUE)

panel_data$GEOID <- sprintf("%012.0f", as.numeric(panel_data$GEOID))
bg$GEOID <- as.character(bg$GEOID)

common_id <- intersect(bg$GEOID, panel_data$GEOID)
bg2 <- bg[bg$GEOID %in% common_id, ]
```

```
nb <- poly2nb(bg2, row.names = bg2$GEOID)
W  <- nb2mat(nb, style = "B", zero.policy = TRUE)

##Shape
car_mod_SHAPE <- fitme(
  Join_Count ~ scale(SHAPE_AM) + Per_OlderAdults + Per_Female + MedianIncome_z +
    Per_Poverty + Per_White + Per_BachelorOrHigher + factor(year) +
    adjacency(1 | GEOID) + offset(log(Population)),
  adjMatrix = W,
  data = panel_data,
  family = "negbin1"
)

summary(car_mod_SHAPE)

coefs <- as.data.frame(summary(car_mod_SHAPE)$beta_table)

coefs$IRR <- exp(coefs$Estimate)
coefs$Lower <- exp(coefs$Estimate - 1.96*coefs$`Cond. SE`)
coefs$Upper <- exp(coefs$Estimate + 1.96*coefs$`Cond. SE`)
coefs <-coefs[-1,]

coefs
```
